# Dynamics of *ORF1ab* and *N* Gene among hospitalized COVID-19 positive cohorts: A hospital based retrospective study

**DOI:** 10.1101/2020.11.22.20236240

**Authors:** Pojul Loying, Vaishali Sarma, Suranjana C. Hazarika, Monjuri Kataki, Dina Raja, Divyashree Medhi, Ridip Dutta, Achu Chena, Divya Daimary, Aakangkhita Choudhury, Lahari Saikia

## Abstract

1.

**Objective:** The present study hospital based retrospective study aimed at investigating the dynamics of ORF1ab and N gene from hospitalized COVID-19 positive cohorts considering the Ct values of both genes.

**Study design and Methodology:** Retrospective analyses of Ct values were done from 115 hospitalized COVID-19 positive patients in different time interval. Patients were admitted to the hospital either by RAT or/and RT-PCR and first RT-PCR testing were made after 9 days of incubation followed by testing in every 3 days of interval till negative, subsequently release of the patients.

**Results:** We have looked into the dynamics of ORF1ab and N gene and found that N gene require longer duration of days with 12.68 (S.D.±3.24) to become negative than ORF1ab with 12.09 (S.D.±2.88) days and it differs significantly (p=0.012; p<0.05). The persistent of N gene found in 46 patients out of 115 (39.65%) to the succeeding reading after 3 days. We have also looked into the mean differences in the between N and ORF1ab genes every readings separately and found that there were no significant differences between the mean Ct value of ORF1ab and N gene except in the day 3 (p=0.015; p<0.05). Further, we have looked into the relationship of age and gender of patients with the duration of positivity; however we did not find any significant role.

**Conclusion:** In COVID-19 hospital positive cohorts, the persistent of positivity of N gene is significantly for more duration than ORF1ab. As the SARS-CoV-2 is a new virus and study on it is evolving, so, exhaustive study is required on the dynamic of N gene positivity persistent in relation to the other pathophysiological parameters for the management and control of COVID-19.

## 2. Introduction

A novel Coronavirus, known as Severe Acute Respiratory Syndrome Coronavirus-2 (SARS-CoV-2), initially originated from Wuhan, Hubei province, China in December 2019, wobbled the entire world causing a pandemic disease called Coronavirus-2019 (COVID-19) become a global concerned and disruptive health hazard [1-3]. As of 16^th^ October 2020 (13:30 IST), more than 39.1 million of the world population confirmed positive for SARS-CoV-2 infection with more than 1.1 million death and 1% population of active cases are in serious or critical conditions [4]. India, leading the second position after USA with more than 7.3 million confirmed positive cases with 0.11 million death and positive cases still surging exponentially which is a matter of concern [5]. SARS-CoV-2 is a positive strand RNA virus with genome size of 30,000 bases in length and is belongs to *Betacoronavirus* family shares 79% and 50% genome sequence identity with previous epidemic viruses of this family, SARS-CoV and MERS-CoV respectively [2, 6-8]. The virus is characterized by a unique combination of polybasic cleavage sites, a distinctive feature known to increase pathogenicity and transmissibility in other viruses [6, 8]. Despite of all efforts from scientific community across the globe, till date, no such convincing strategy to cure COVID-19 is discovered either in the form of drug or vaccine; however its control is solely depends on management level, be it repurpose drugs or social measures.

Reverse Transcriptase Real Time Polymerase Chain Reaction (rtRT-PCR) is a gold standard molecular diagnostic tool to detect wide range of virus due to its rapidity, sensitivity, reproducibility and the reduced risk of contamination and it has become an essential tool for the confirmation of COVID-19 disease by detecting the presence of SARS-CoV-2 from the upper respiratory (nasopharyngeal and oropharyngeal) swab samples [9-11]. Since the first availability whole genome sequencing from Wuhan, many target gene sequences identified for the detection of SARS-CoV-2 genome by using RT-PCR and the conserve domains including *N* gene (Nucleocaspid), *RdRp* gene (RNA dependent RNA polymerase), *E* gene (Envelope), *S* gene (Spike), *H* gene (Helicase), *HE* gene (Hemagglutinin-esterase) and ORF (Open reading frame) group of genes [7, 12-16]. More than two third of the SARS-CoV-2 genome encoded by *ORF1ab* gene of 21,290 nucleotides at the 5’ end and in addition 6 other ORF genes including *ORF3a, ORF6, ORF7a, ORF7b*, and *ORF8* genes [7]. Nucleocapsid, a type of structural protein of SARS-CoV-2 is encoded by 908 nucleotides *N* gene at the near 3’ end and it’s a potential target for the detection of virus as recommended by CDC, United States [7, 17]. It is always advisable to use two molecular targets at a time to avoid potential cross- reaction with other endemic coronaviruses as well as potential genetic drift of SARS-CoV-2 [18, 19]. Studies from many countries reveal that investigators adopting two target strategy like Berlin Institute of Virology, Berlin and German Centre for Infection Research (DZIF), Berlin, Germany used *RdRp* and *E* gene [16]; National Institute for Viral Disease Control, China CDC used *ORF1ab* and *N* gene [20]; CDC, United States used two loci of *N* gene (*N1* and *N2*) [17].

Threshold cycle (Ct) value of a target gene in RT-PCR based viral molecular diagnosis not only gives information about the presence or absence of a virus, apart it also helps in understanding the dynamics of a virus in due course of infection; however, clinical importance is limited due to various laboratory complications [21, 22]. The Ct value is defined as the cycle number when the sample fluorescence exceeds a chosen threshold and signifies by lower the Ct value of a specific gene, the more the gene exists in the sample [22, 23]. Ct values are therefore inversely related to viral load and can provide an indirect method of quantifying the copy number of viral RNA in the sample; eventually the viral load [24]. Viral dynamics knowledge is essential for formulating strategies for treatment, management and epidemiological control of COVID-19 [25]. In a cohort study by Chen, Y. *et al*. on oropharyngeal saliva samples shown the dynamics where salivary viral load was highest during the first week after symptom onset and subsequently declined with time [26]. SARS-CoV-2 viral load in upper respiratory specimens of infected patients observed the progressive decrease of viral load over time with positivity persist till 17–21 days after onset of symptoms [27]. Many studies on viral dynamics of COVID-19 positive cohorts evidenced that Ct value of a target gene is inversely proportional to viral loads irrespective of patient’s conditions; however several technical problems need to be considered from sample collection to RT-PCR testing [28-32].

Till date, no such comprehensive study on the dynamics of virus in hospitalized COVID-19 patients in different time point, till negative and discharge has been made. In the present study, we are exhaustively exploring the dynamics of *ORF1ab* and *N* gene from the 115 (one hundred and fifteen) hospitalized COVID-19 positive cohorts of India using different time interval, starting from the first RT-PCR test to the negative, eventually discharged.

## 3. Materials and Methods

### 3.1. Study design

The present study was carried out to characterize the dynamics of *ORF1ab* and *N* gene in 115 hospitalized COVID-19 positive patients at Gauhati Medical College and Hospital (GMCH), Assam, India. The hospital is one among the major and largest in COVID-19 testing and treatment in eastern India. Only two parameters; age and gender of patients were considered for the study. Any other parameters like source of infection, patient conditions, treatment regime and other comorbidities were not considered for this study. The COVID-19 positive confirmation was made as per the guidelines of Indian Council of Medical Research (ICMR), Govt. of India. The RT-PCR profiling was performed for the hospitalized COVID-19 positive patients at regular time interval and Ct values were recorded from the day of first testing in the hospital till negative; subsequently discharge of the patients. The patients were hospitalized after being tested positive either by Rapid Antigen Test or RT-PCR. All the patients were kept for 9 (nine) days incubation followed by first tests and subsequently tests after 3 (three) days of interval till negative. The day 9, day of first testing was taken as 0 (zero) day; technically the days of testing are considered as (9+n); where n is 0, 3, 6… days **(Figure 1)**. The Ct values of RT-PCR results were considered for study and all the negative results were taken as Ct value of 40 for the convenient of the study. The RT-PCR data were collected in the month of September, 2020. RT-PCR data were taken for analysis after approval from institutional ethical committee (IEC), GMCH. Consent from the positive patients was waived due to the retrospective nature of the analysis.

**Figure 1:**
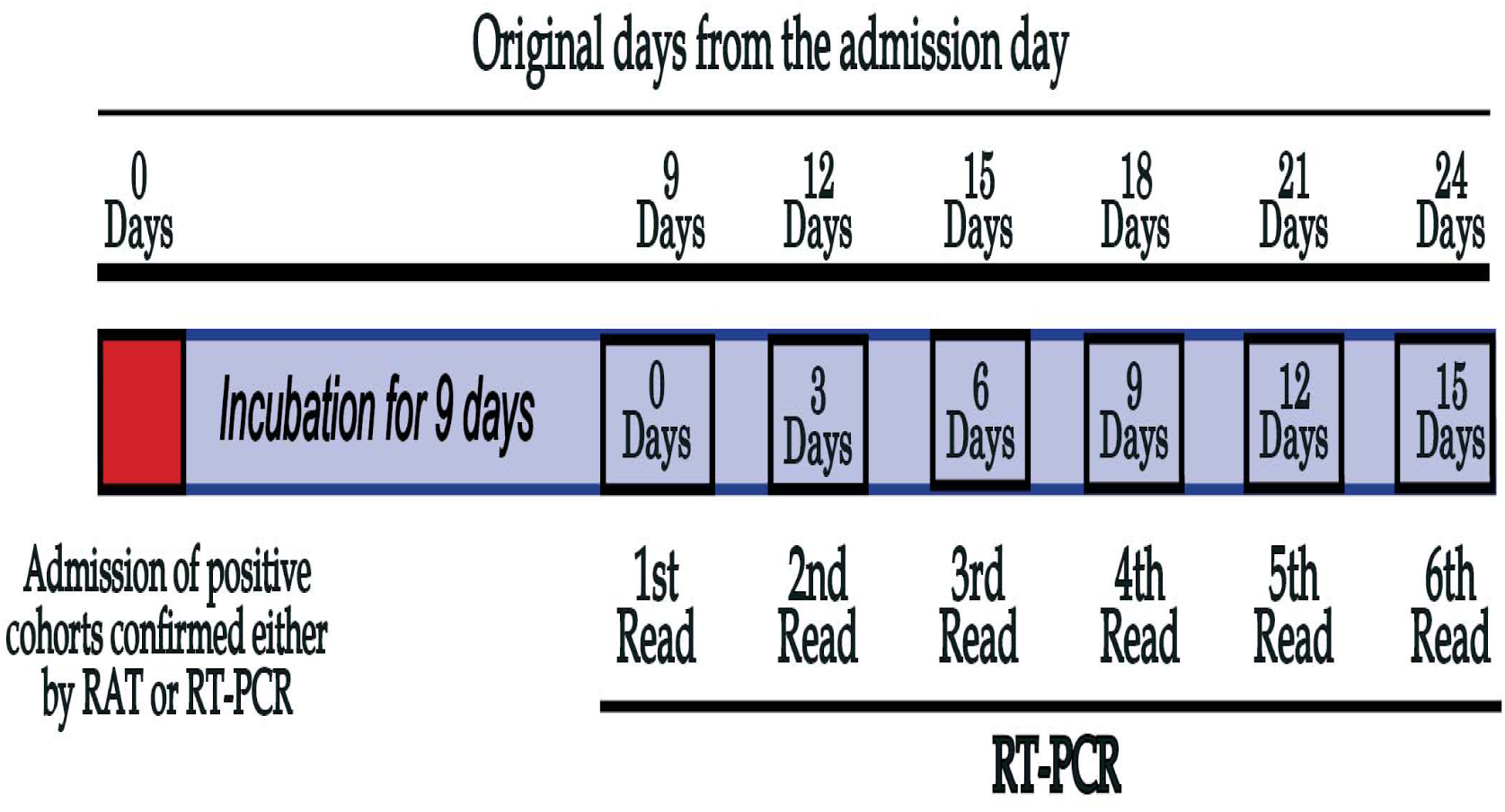
Overall strategy for RT-PCR test of hospitalized COVID-19 positive patients.

### 3.2. Sample collection, processing and RNA extraction

All the kits used for the present study were approved by Indian Council for Medical Research (ICMR), Govt. of India. Throat swab and deep nasal swab samples were collected for SARS-CoV-2 detection from hospitalized COVID-19 positive patients by using sterile swab stick and subsequently both swabs were put in ViSure Transport Kit (Labsystems Diagnostics, Finland). The samples were further processed in laboratory by vortex and aliquots were made in 1.5 ml MCT. RNA from the processed samples was extracted by using QIAmp Viral RNA Mini Kit (QIAGEN, Germany) following manufacturer guidelines.

### 3.3. RT-PCR for SARS-CoV-2 detection

One Step Reverse Transcriptase Real Time Polymerase Chain Reaction (RT-PCR) were performed to detect the presence or absence of *ORF1ab* and *N* gene using Taqman probe based multicolor Meril COVID-19 One-step RTPCR Kit (Meril Diagnostics, Belgium) in CFX96 Touch Real-Time PCR Detection System RT-PCR platform (Bio-Rad Laboratories, Inc.). Meril COVID-19 One-step RTPCR Kit is approved by Drug Controller General of India (DCGI), Food and Drug Administration (FDA) and Foundation for Innovative New Diagnostics (FIND), Govt. Of India as RT-PCR molecular test that can be used in patient care settings [19]. Only single kit was considered for the experiment due kit to kit variations and issue of efficiency. The reaction mixture preparation and amplification program was used as per manufacturer recommendation. The result interpretation was made according to manufacturer instruction considering threshold cycle value (Ct≤40) for both *ORF1ab* & *N* genes and Internal Control (IC).

### 3.4. Statistical analysis

Continuous variables were first tested for normality and homogeneity of variance, and accordingly proceed with the succeeding tests. The continuous variables with two groups were tested using Welch Two Sample t-test or Mann-Whitney U test. Continuous variables with more than two groups were tested using Kruskal-Wallis rank sum test followed by Dunn’s Multiple Comparision test to compare the differences between the groups. Categorical values were tested using Chi-Square test for Independence. For all the tests, P-Values less than 0.05 were considered as statistically significant. All statistical analyses were performed in R (version 4.0.0) and SPSS (version 16.0) software. The graphical representations were created in R software (version 4.0.0) using GGPLOT2 package.

## 4. Results and Discussion

To study the dynamics of *ORF1ab* and *N* gene in hospitalized COVID-19 positive cohorts from nasopharyngeal and oropharyngeal swab, we have considered 115 patients from an Indian COVID-19 hospital for the present retrospect study.

Our study shows that the Ct value gradually increases (tends to 40) with time in the succeeding readings **(Figure 2a)**. We have also considered Ct value ≥40 as negative for SARS-CoV-2. Further, we have observed that the *ORF1ab* and *N* gene positivity of patients also decreases with time i.e. in every succeeding RT-PCR **(Figure 2b)**. This changes in mean Ct values with succeeding days was found significant for both the genes (p=0.01; p<0.05); however, in post hoc analysis it differs **(Table 1)**. Low Ct value in RT-PCR is considered as high viral load and vice versa, though various experimental pitfalls have to consider [21, 33]. Few studies on dynamics of SARS-CoV-2 viral load evidenced that higher Ct value correlate with low viral load and Ct value or viral load has somehow correlation with symptoms, disease progression, disease severity and death [24, 28, 30, 32, 34]. La Scola B. *et al* cultured SARS-CoV-2 on Vero cell line and show that there was a clear correlation between Ct value and viral load in culture. In the study, they found growth in culture from the patients with Ct≤34; however, no growth observed in Ct more than 34 and patients were discharged [35].

**Table 1:**
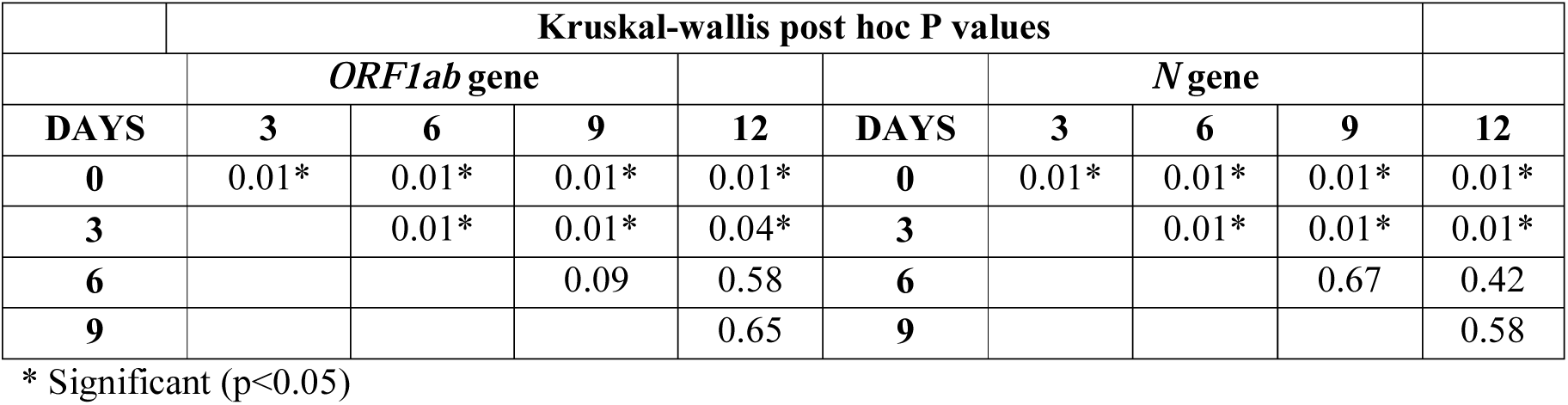
Post hoc test to show the changes in mean Ct values with succeeding days.

**Figure 2:**
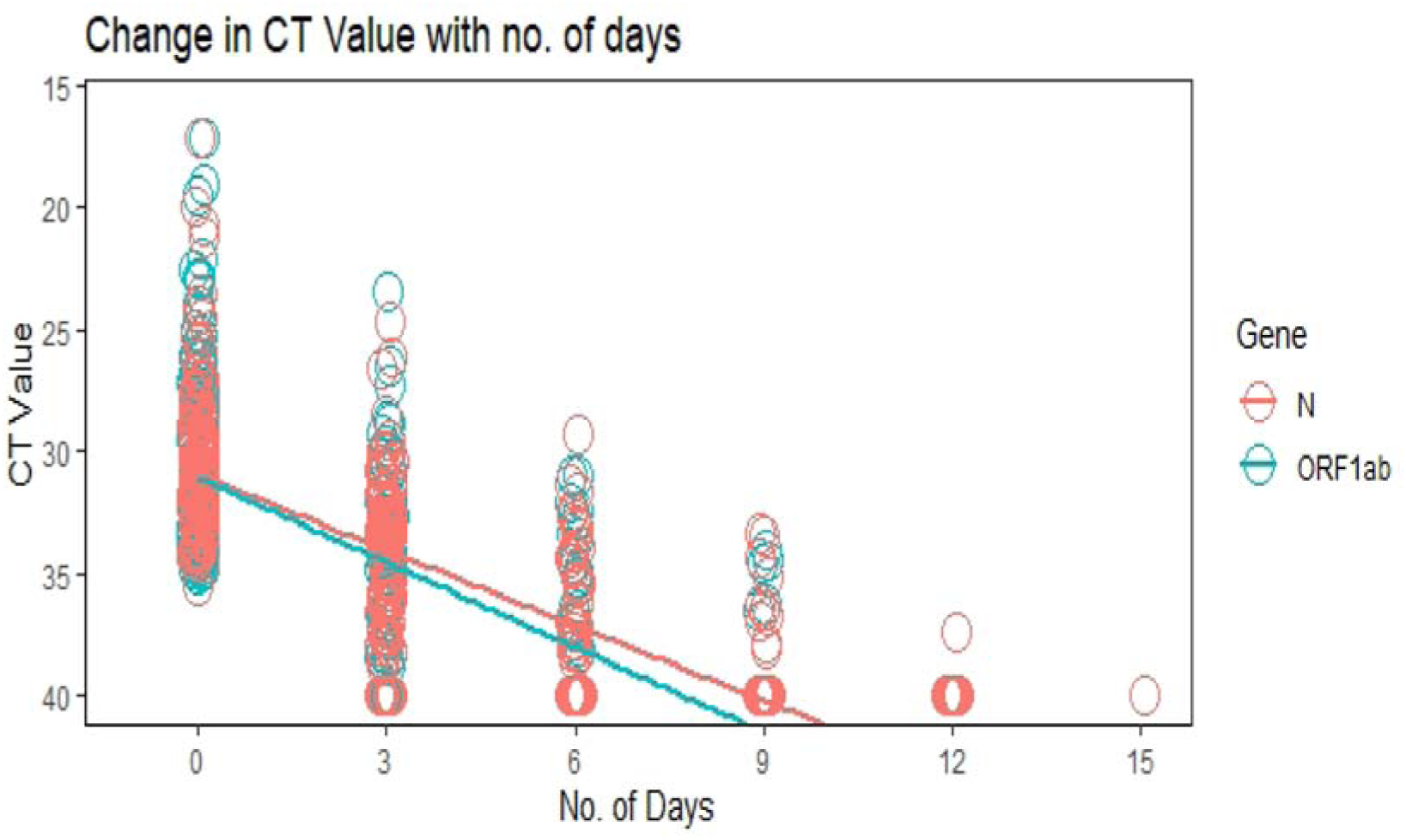
Distribution of Ct value of *ORF1ab* and *N* gene vs. RT-PCR reading at different time point

**Figure 2b:**
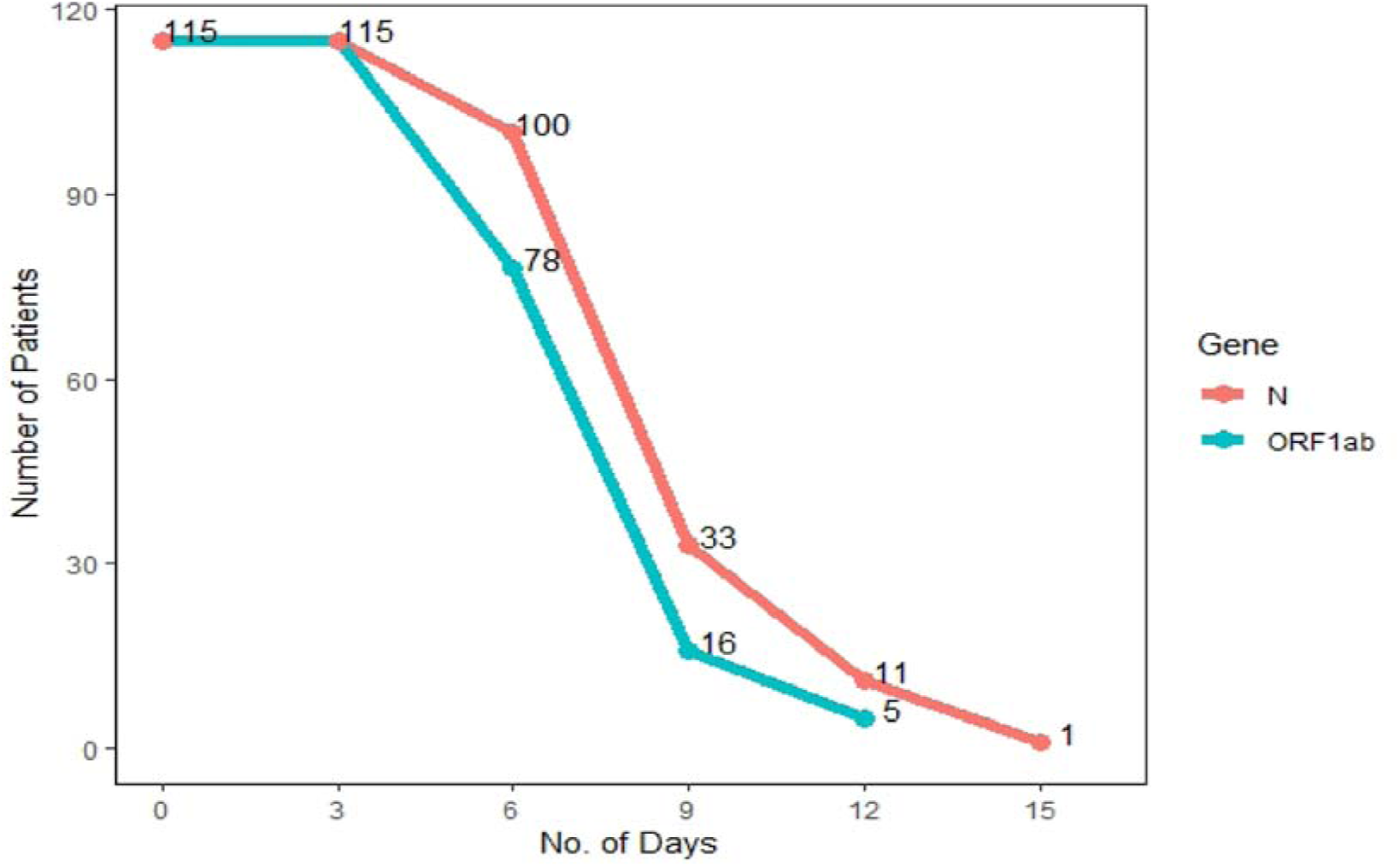
Distribution of *ORF1ab* and *N* gene of patients in every succeeding RT-PCR readings.

Our dynamic study show that *ORF1ab* gene required less time to become negative with mean value of 3.09 (S.D. ± 2.88) days to become negative from the day of 1st testing; where overall days from the day of hospital admission was (9+n), where ‘9’ is the incubation days common for all and ‘n’ is the number of days after 1st testing i.e. 12.09 (S.D.±2.88) days than *N* gene. For *N* gene, mean days to become negative is 3.68 (S.D. ±3.24) from the 1st testing with overall days of 12.68 (S.D.±3.24). The overall range of days for negative for *ORF1ab* is 12 to 21 and *N* gene is 12 to 25. Comparison between the mean days of two genes using Welch’s t-test showed the persistent of *N* gene is significantly for longer period than *ORF1ab* gene (p=0.012; p<0.05) **(Figure 3)**. The persistent of *N* gene found in 46 patients out of 115 (39.65%) to the succeeding reading after 3 days. In a study on the correlation between Ct value of *N* gene show that persistent is significantly for longer period in female than male; however, there was an strong correlation observed in between male and persistence of virus for longer period [34]. In a preprint article, a study done on Moroccan hospitalized positive cohorts show than *N* gene persist for long days than *RdRp* and *E* gene in 3% of study populations [36]. However, the exact mechanism of the persistent of *N* gene in nasopharyngeal and throat swab for longer is not well studied. In an extensive review on Coronavirus N protein by McBride R, *et al* explained that the N protein is abundantly produced within infected cells and perform multiple functions including binding to viral RNA to form the ribonucleocapsid and has also been proposed to have roles in virus replication, transcription and translation [37]. In an extensive study by Wolfel R, *et al* explained that that positive detection of SARS-CoV-2 reflects the presence of RNA, not necessarily viable [38].From the above information, we can assume that persistent of *N* gene for longer duration, may be a detection from our own infected cells, not from virus; as no growth observed in cell culture in Ct more than 34 by La Scola, *et al*, or from nonviable SARS-CoV-2; however an extensive study is must need [35, 38].

**Figure 3:**
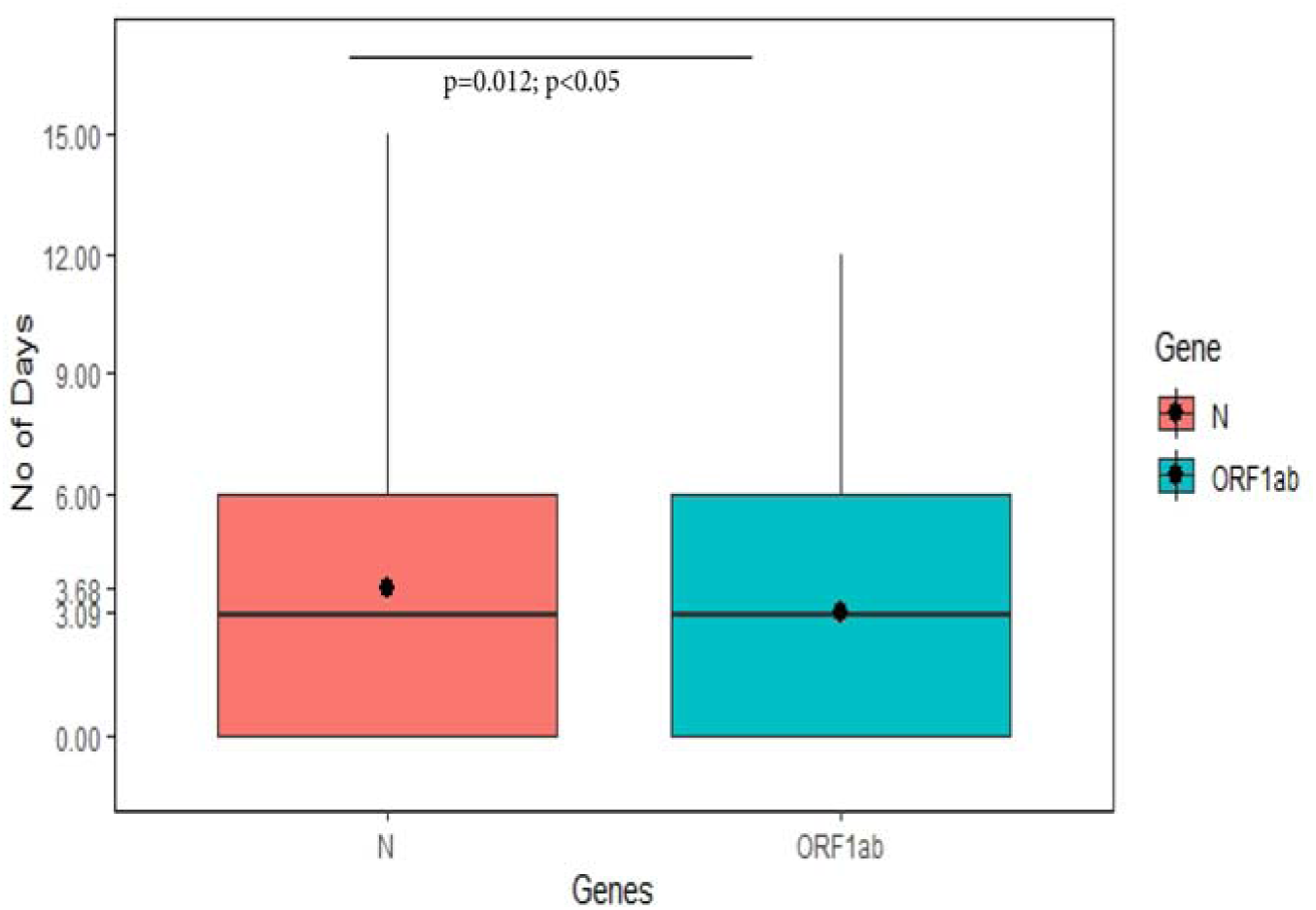
Differences in mean duration of days between *ORF1ab* and *N* gene

One may argue that the persistent of *N* gene for longer period may arise due to sensitivity of *N* over *ORF1ab*. To address this question we have looked into the mean differences in the between *N* and *ORF1ab* genes every readings separately. We found that there were no significant differences between the mean Ct value of *ORF1ab* and *N* gene except in the day 3 (p=0.015; p<0.05) **(Figure 4) (Table 2)**. The particular finding nullified the issue of sensitivity of two genes. Further, we looked into the relationship of age and gender with the duration of positivity and we found that there was no significant role of age and gender with the positivity duration.

**Table 2:**
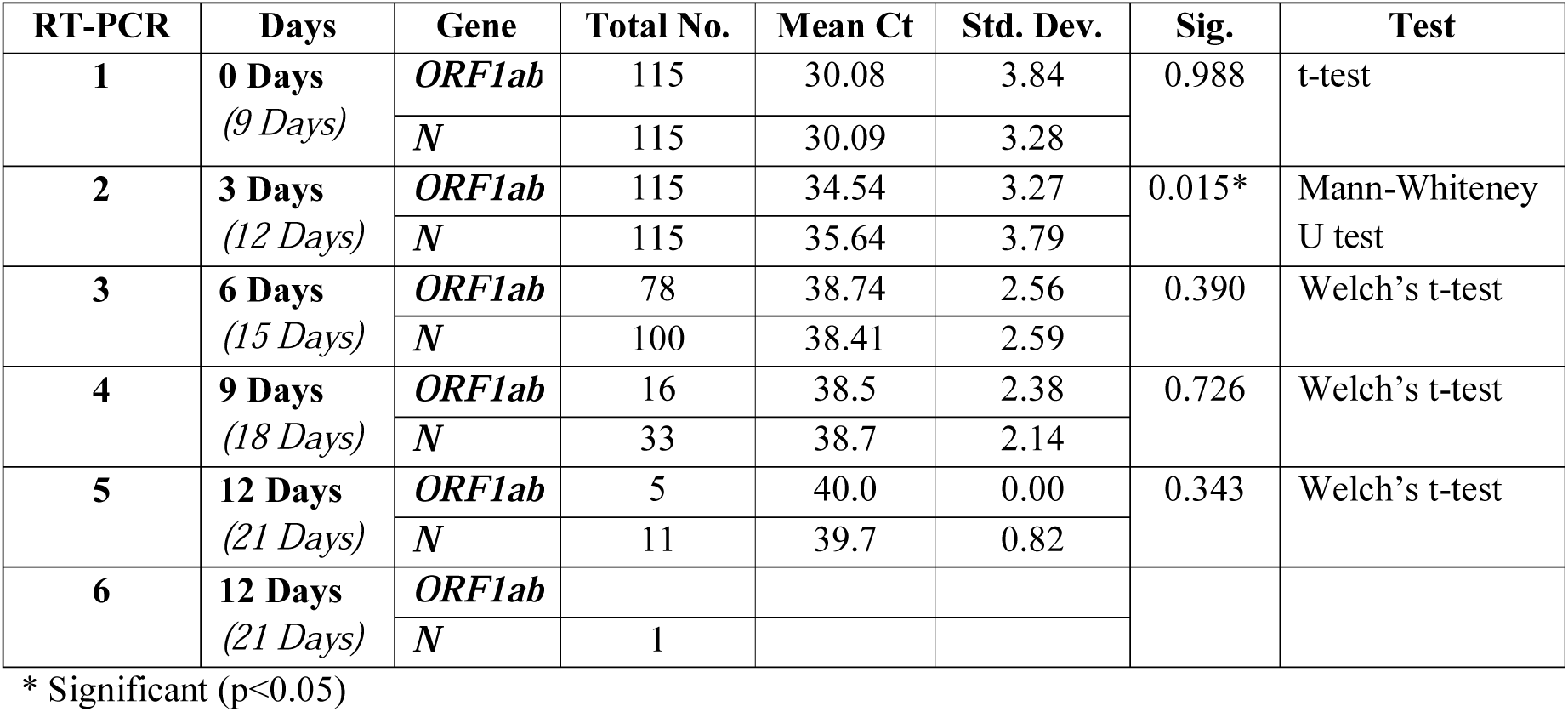
Mean Ct value of RT-PCR in different time point with other statistical parameters.

| **RT-PCR** | **Days** | **Gene** | **Total No.** | **Mean Ct** | **Std. Dev.** | **Sig.** | **Test** |
| --- | --- | --- | --- | --- | --- | --- | --- |
| **1** | **0 Days**<br>(9 Days) | *ORF1ab* | 115 | 30.08 | 3.84 | 0.988 | t-test |
|  |  | *N* | 115 | 30.09 | 3.28 |  |  |
| **2** | **3 Days**<br>(12 Days) | *ORF1ab* | 115 | 34.54 | 3.27 | 0.015* | Mann-Whitney<br>U test |
|  |  | *N* | 115 | 35.64 | 3.79 |  |  |
| **3** | **6 Days**<br>(15 Days) | *ORF1ab* | 78 | 38.74 | 2.56 | 0.390 | Welch's t-test |
|  |  | *N* | 100 | 38.41 | 2.59 |  |  |
| **4** | **9 Days**<br>(18 Days) | *ORF1ab* | 16 | 38.5 | 2.38 | 0.726 | Welch's t-test |
|  |  | *N* | 33 | 38.7 | 2.14 |  |  |
| **5** | **12 Days**<br>(21 Days) | *ORF1ab* | 5 | 40.0 | 0.00 | 0.343 | Welch's t-test |
|  |  | *N* | 11 | 39.7 | 0.82 |  |  |
| **6** | **12 Days**<br>(21 Days) | *ORF1ab* |  |  |  |  |  |
|  |  | *N* | 1 |  |  |  |  |

\* Significant (p<0.05)

**Figure 4:**
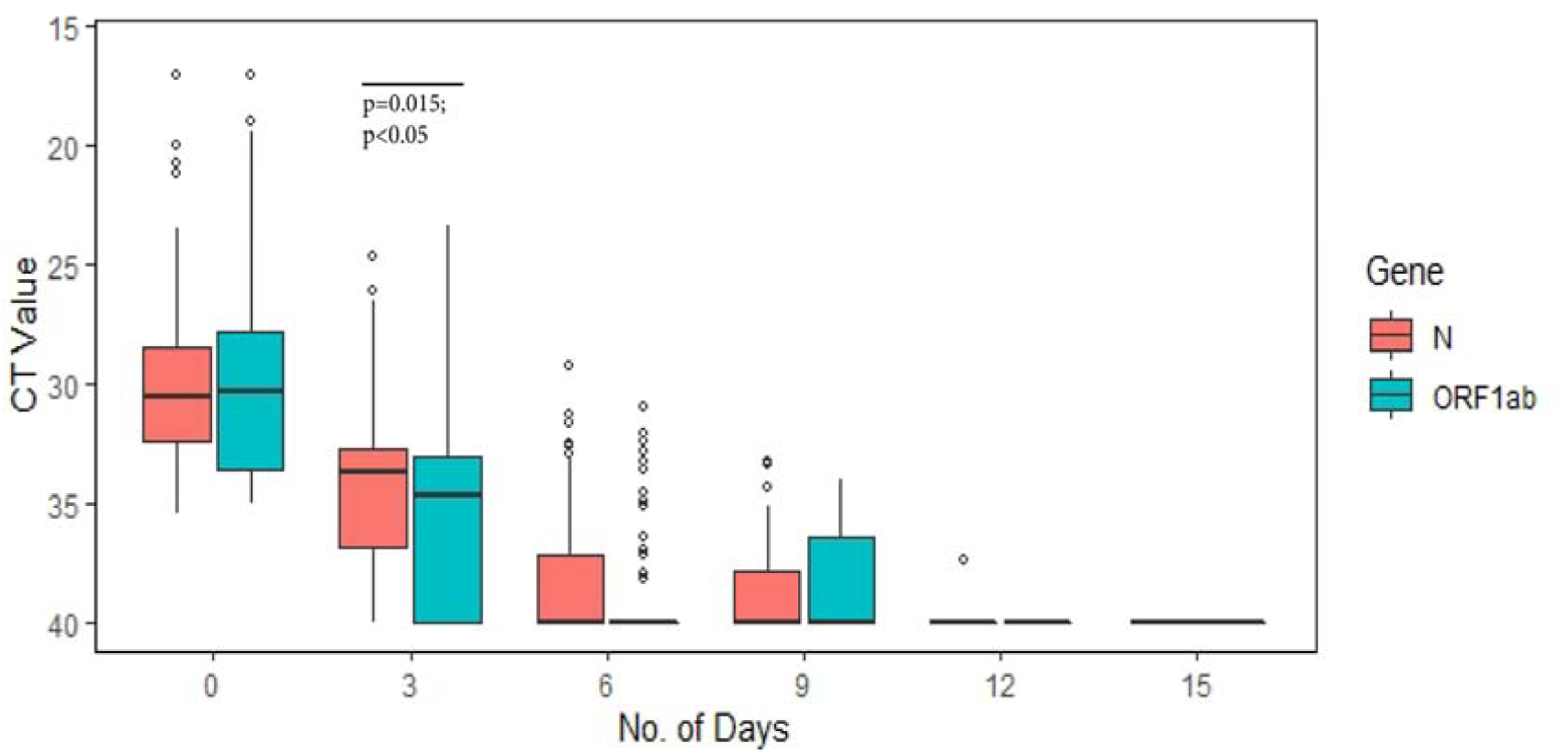
Difference between mean Ct value of *ORF1ab* and *N* gene in different time point

## 5. Conclusion

In this work, we have shown the dynamics of *ORF1ab* and *N* gene from the oro- and nasopharyngeal swab of hospitalized COVID-19 positive cohorts, where the mean day duration required to become negative for *N* gene with 12.68 (S.D.±3.24) is significantly longer than the *ORF1ab* gene 12.09 (S.D.±2.88). In 46 out of 115 (39.65%) patients, the persistent of *N* gene was found, with maximum days required for negative for *N* gene is for 25 days whereas 21 days for *ORF1ab* gene. Further, we have looked into the relationship of age and gender of patients with the duration of positivity; however we did not find any significant role.

As the study on SARS-CoV-2 is evolving, there is a lack of knowledge on viral gene dynamics, especially persistent of N gene that we have shown, and correlation of persistent with the pathogenesis and pathophysiology of this novel virus infection. Exhaustive study on the persistent of *N* gene will give more information in the control and management of this pandemic COVID-19.

## Data Availability

Data availabe within the article

## 7. Acknowledgement

Authors acknowledge Puja Bishaya, a PhD student, Cotton University, Assam, India for her proactive help in statistical analysis.

